# Characterizing responsiveness to the COVID-19 pandemic in the United States and Canada using mobility data

**DOI:** 10.1101/2022.11.08.22282050

**Authors:** Jean-Paul R. Soucy, David N. Fisman, Derek R. MacFadden, Kevin A. Brown

## Abstract

**Background:** Mobile phone-derived human mobility data are a proxy for disease transmission risk and have proven useful during the COVID-19 pandemic for forecasting cases and evaluating interventions. We propose a novel metric using mobility data to characterize responsiveness to rising case rates.

**Methods:** We examined weekly reported COVID-19 incidence and retail and recreation mobility from Google Community Mobility Reports for 50 U.S. states and nine Canadian provinces from December 2020 to November 2021. For each jurisdiction, we calculated the responsiveness of mobility to COVID-19 incidence when cases were rising. Responsiveness across countries was summarized using subgroup meta-analysis. We also calculated the correlation between the responsiveness metric and the reported COVID-19 death rate during the study period.

**Findings:** Responsiveness in Canadian provinces (*β* = -1·45; 95% CI: -2·45, -0·44) was approximately five times greater than in U.S. states (*β* = -0·30; 95% CI: -0·38, -0·21). Greater responsiveness was moderately correlated with a lower reported COVID-19 death rate during the study period (Spearman’s *ρ* = 0·51), whereas average mobility was only weakly correlated the COVID-19 death rate (Spearman’s *ρ* = 0·20).

**Interpretation:** Our study used a novel mobility-derived metric to reveal a near-universal phenomenon of reductions in mobility subsequent to rising COVID-19 incidence across 59 states and provinces of the U.S. and Canada, while also highlighting the different public health approaches taken by the two countries.

**Funding:** This study received no funding.

**Research in context:** *Evidence before the study:* There exists a wide body of literature establishing the usefulness of mobile phone-derived human mobility data for forecasting cases and other metrics during the COVID-19 pandemic. We performed a literature search to identify studies examining the opposite relationship, attempting to quantify the responsiveness of human mobility to changes in COVID-19 incidence. We searched PubMed on October 21, 2022 using the keywords “COVID-19”, “2019-nCoV”, or “SARS-CoV-2” in combination with “responsiveness” and one or more of “mobility”, “distancing”, “lockdown”, and “non-pharmaceutical interventions”. We scanned 46 published studies and found one that used a mobile phone data-derived index to measure the intensity of social distancing in U.S. counties from January 2020 to January 2021. The authors of this study found that an increase in cases in the last 7 days was associated with an increase in the intensity of social distancing, and that this effect was larger during periods of lockdown/shop closures.

*Added value of the study:* Our study developed a metric of the responsiveness of mobility to rising case rates for COVID-19 and calculated it for 59 subnational jurisdictions in the United States and Canada. While nearly all jurisdictions displayed some degree of responsiveness, average responsiveness in Canada was nearly five times greater than in the United States. Responsiveness was moderately associated with the reported COVID-19 death rate during the study period, such that jurisdictions with greater responsiveness had lower death rates, and was more strongly associated with death rates than average mobility in a jurisdiction.

*Implications of all the available evidence:* Mobile phone-derived human mobility data has proven useful in the context of infectious disease surveillance during the COVID-19 pandemic, such as for forecasting cases and evaluating non-pharmaceutical interventions. In our study, we derived a metric of responsiveness to show that mobility data may be used to track the efficiency of public health responses as the pandemic evolves. This responsiveness metric was also correlated with reported COVID-19 death rates during the study period. Together, these results demonstrate the usefulness of mobility data for making broad characterizations of public health responses across jurisdictions during the COVID-19 pandemic and reinforce the value of mobility data as an infectious disease surveillance tool for answering present and future threats.

## Introduction

Mobile phone-derived human mobility data have proven to be an invaluable data source for infectious disease surveillance during the COVID-19 pandemic. Since the early days of this global disease outbreak, companies including Google, Apple, and SafeGraph have provided datasets for understanding patterns of human mixing and movement and ultimately disease transmission.^1,2^ Researchers have used mobility data for estimating the effectiveness of non-pharmaceutical interventions^3,4^, forecasting incident cases^5–9^, and to assess the population-level changes required to arrest SARS-CoV-2 transmission.^10^

Much attention has been paid to mobility as a predictor of COVID-19 incidence and changes in mobility as a response to clearly defined non-pharmaceutical interventions such as lockdowns and other physical distancing measures. However, changes in mobility are also driven by organic, individual-level responses.^11^ For example, public transit use in March 2020 declined in major cities across the globe prior to the official imposition of public health measures in their respective countries, likely driven by media coverage and personal risk assessment.^12^ Thus, aggregated mobile phone data reflect both top-down policy prescriptions and bottom-up population responses to perceived COVID-19 risk.

Throughout much of the pandemic, the most accessible gauge of COVID-19 risk was the reported rate of new COVID-19 cases. This quantity was often presented in the context of an epidemic curve, which also shows whether cases are rising or falling. Changes in human mobility following high or rising reported case rates can thus be thought of as an indicator of population-level “responsiveness” to the pandemic, whether this is achieved through government policy, individual decision making based on personal risk assessment, or a combination of both.

In this analysis, we characterized the responsiveness of population-level mobility to reported levels of COVID-19 incidence in 59 states and provinces across the United States and Canada between December 2020 and November 2021.

## Methods

### Study design and population

This ecological study used meta-analysis to contrast the responsiveness of population-level mobility to reported COVID-19 incidence in 59 subnational jurisdictions across the United States and Canada, comprising all 50 US states and nine out of ten Canadian provinces. The province of Prince Edward Island and the three Canadian territories were excluded because their small population sizes rendered their mobility data unreliable.

This study covered the 12-month period between December 2020 and November 2021. We chose this period for several reasons. First, it spans a single, full seasonal cycle, which is important since the seasonality of the disease differs across jurisdictions included in the study. Second, the availability of confirmatory testing for SARS-CoV-2 infection was generally adequate across this time period. Third, this period begins at the cusp of both the vaccine roll-out and the first variant-driven wave (caused by the Alpha variant), after which followed strong differentiation in pandemic response measures between jurisdictions. Finally, this period concludes just prior to the dominance of the Omicron variant, which resulted in the availability of confirmatory testing being severely curtailed in some jurisdictions.

Data were aggregated to the level of week to remove day-of-the-week effects, so the study comprised the 52 weeks beginning with the week of December 6, 2020 and ending with the week of November 28, 2021.

This project was approved by the University of Toronto Research Ethics Board (RIS Human Protocol Number 43389).

### Measurement of outcome

The outcome, weekly change in mobility, was measured using the state/province-level “retail and recreation” metric of Google’s Community Mobility Reports.^13,14^ These mobility reports measure, for various levels of geography, the relative number of visits to five categories of places: retail and recreation, grocery and pharmacy, parks, transit stations, and workplaces compared to a baseline period (January 3 to February 6, 2020). It also includes the relative number of hours spent at home (residential) compared to the same baseline period. For example, a value of 100% for the retail and recreation metric indicates the same volume of visits to these types of locations as during the baseline period; a value of 90% indicates that the volume of visits is 10% lower than during the baseline period.

We chose retail and recreation over other mobility metrics for three reasons. First, retail and recreation activities are demonstrably highly responsive to pandemic-related measures (unlike grocery stores and pharmacies, which sell goods essential to the whole population).^15^ Second, we expect the metric to be more representative of whole-population mobility compared to workplaces (reflecting in-person workers) and transit stations (reflecting primarily urban residents). Finally, we expected it to be less seasonal than some metrics (especially park visits and time spent at home).

Highly seasonal metrics would be expected to change significantly through the year, compared to the early 2020 baseline period, irrespective of the changing pandemic situation.

We defined the outcome of our analysis, the weekly *change* in mobility, as the difference in the average value of the mobility metric in the current week compared to the average value in the prior week (for example, mobility going from 100% of baseline to 90% of baseline represents a -10% change).

### Measurement of exposure

The reported COVID-19 case rate per 1,000 population in the previous week was measured by summing cases reported to the national public health body by each state/province, dividing by the 2021 mid-year population estimate, and multiplying the result by 1,000. Additional lags (i.e., case rate for two weeks prior and three weeks prior) were also calculated. See supplementary methods for detailed data sources.

### Statistical analysis

We propose a model for measuring responsiveness where the change in mobility in the current week (*Δ*_*mobility*_) is a function of the observed COVID-19 case rate in the previous week. A negative slope implies that mobility decreases when the case rate increases. The more steeply negative the slope, the stronger the implied responsiveness of mobility to observed COVID-19 case rate is.

Additionally, we add two terms to the model: a dummy variable for whether the case rate in the previous week was falling (or staying the same) compared to the week before that (or whether it was rising) and an interaction term between the case rate and cases falling variable. These additional terms allow the intercept and slope to be different for times when cases are rising versus times where cases are falling. This is because, for the same absolute COVID-19 case rate, we might expect measures to be taken to reduce mobility when cases are rising but not when cases are falling. In this analysis, we focus on responsiveness when cases are rising (*β*_*rising*_), as we hypothesized that it would be greater than responsiveness when cases are falling (*β*_*falling*_).

The model is given by the following linear regression equation:

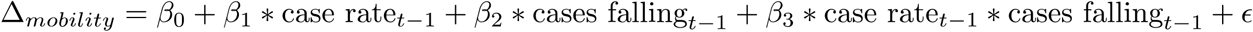

with *Δ*_*mobility*_ measured as mobility at week *t* minus mobility at week *t-1*, case rate measured as the incidence of COVID-19 cases per 1,000 population reported in week *t-1*, and cases falling as a dummy variable with value 0 when the case rate increased from week *t-2* to week *t-1* and 1 when the case rate fell or remained the same from week *t-2* to week *t-1*. Responsiveness when cases are rising (*β*_*rising*_) is given by *β*_*1*_ and responsiveness when cases are falling (*β*_*falling*_) by *β*_*1*_ *+ β*_*3*_.

For each of the 59 jurisdictions in our analysis, we used the above linear regression model to calculate responsiveness. We then used a subgroup meta-analysis approach (random effects inverse variance meta-analysis with restricted maximum likelihood to estimate between-study variance *τ*^*2*^) to compare estimates of responsiveness grouped by country (Canada, United States) or by five region groups defined by Canada and the four U.S. Census Bureau regions (West, South, Northeast, Midwest).

Finally, we calculated the Spearman correlations between *β*_*rising*_ and *β*_*falling*_ and reported COVID-19 death rates during the study period in each jurisdiction. Deaths were retrieved from the same datasets as cases. Deaths were shifted back by four weeks to account for delays between infections and health outcomes as well as reporting delays (i.e., we defined deaths during the study period as those reported between the weeks of 2021-01-03 and 2021-12-26).^16^ For comparison, we also calculated the correlation between the average weekly mobility and death rates during the study period.

We used R (version 4.1.3) for the statistical analysis.^17^

### Sensitivity analyses

We performed two sensitivity analyses.

First, we considered alternative lags for the case rate variable. In addition to considering the case rate at week *t-1*, we fit models looking at the case rate at week *t-2* and week *t-3* (with the corresponding change to the cases falling variable). We hypothesized that the magnitude of responsiveness of mobility to the case rate would be attenuated at longer lags (i.e., *β*_*rising*_ is closer to 0), as more recent information should be given priority for determining population-level mobility.

Second, we considered an alternative linear regression model to estimate responsiveness (interaction-only model):

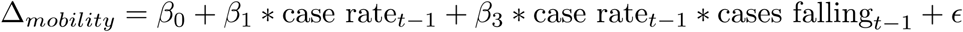

In this alternative model, we remove the term for cases falling (*β*_*2*_), leaving only the interaction term between case rate and cases falling. This allows the slopes to be different when cases are rising versus falling/staying the same, as in the original model, but forces the y-intercept to be the same for both slopes. The justification for this alternative model is that we would not expect there to be much of a difference in the responsiveness of mobility to case rates whether cases are rising are falling when absolute case rates are near 0, resulting in a shared y-intercept for the slopes of *β*_*rising*_ and *β*_*falling*_.

## Results

Reported COVID-19 case rates throughout the study period differed greatly between the United States and Canada, with the U.S. experiencing both much higher average rates and a much higher maximum rate (figure 1).

**Figure 1.**
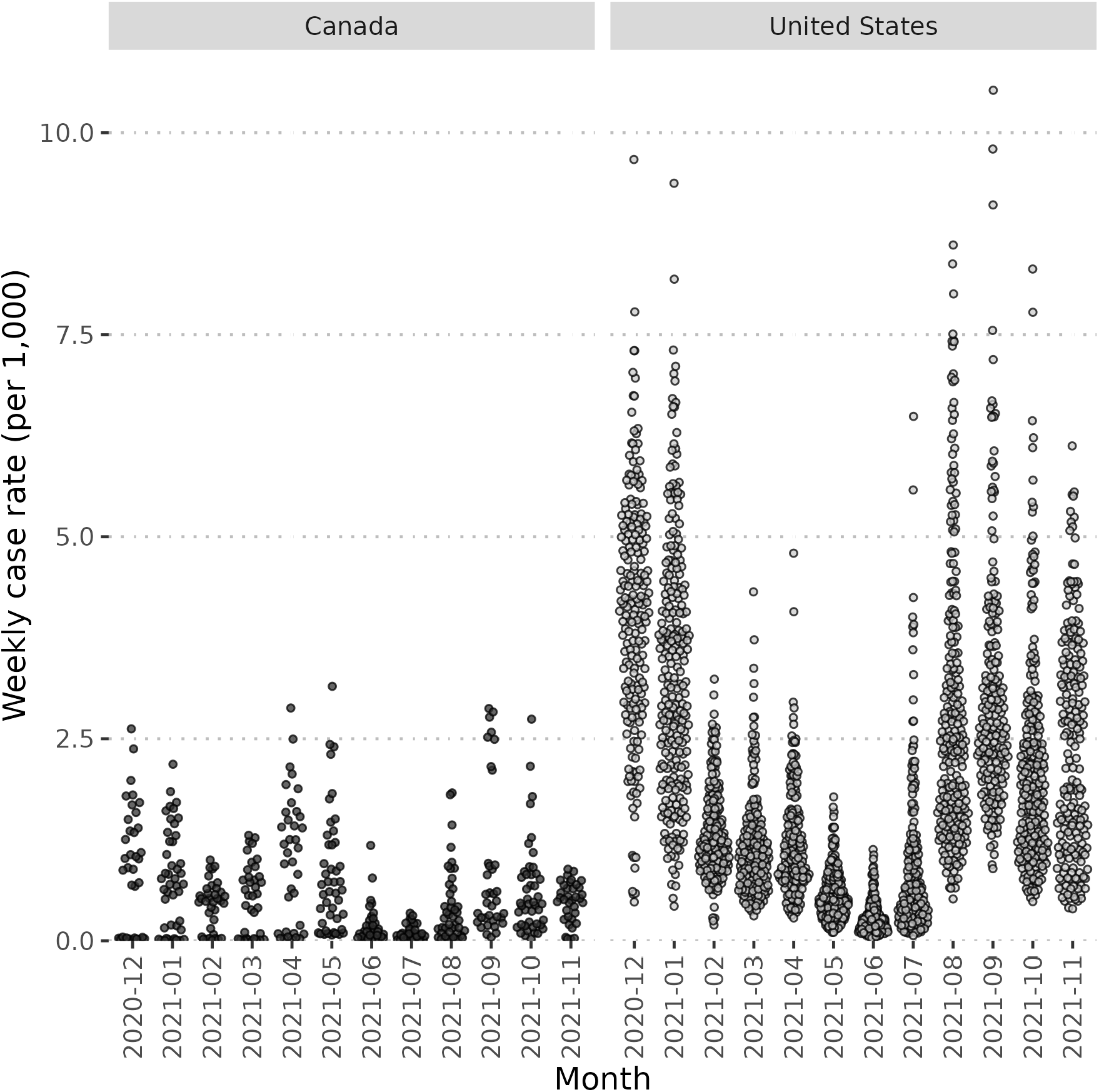
Beeswarm plot of weekly reported COVID-19 case rates per 1,000 population between December 2020 and November 2021 for nine Canadian provinces and 50 US states.

Responsiveness, interpretable as the % change in weekly mobility for a 1 per 1,000 population increase in weekly COVID-19 incidence when cases were rising (*β*_*rising*_), was nearly five times greater on average in Canada (-1·45; 95% CI: -2·45, -0·44) compared to the United States (-0·30; 95% CI: -0·38, -0·21) (table 1, figure 2). The jurisdictions with the highest responsiveness were the provinces of Atlantic Canada (Newfoundland, New Brunswick, Nova Scotia), Ontario, and Quebec, whereas the least responsive jurisdictions tended to be in the South U.S. Census region. (table 1, figure 2). Responsiveness was associated with COVID-19 death rates during the study period (Spearman’s *ρ* = 0·51) (figure 3).

**Table 1.** Pooled beta coefficients for the association between weekly COVID-19 case rates and changes in mobility in the subsequent week when cases are rising (*β*_*rising*_) from subgroup meta-analysis by country and region groups.

| **Subgroup** | **Beta coefficient (95% CI)** |
| --- | --- |
| Overall | -0.31 (-0.39, -0.22) |
| Countries |  |
| Canada | -1.45 (-2.45, -0.44) |
| United States | -0.30 (-0.38, -0.21) |
| Test for subgroup differences (*p*) | 0.0258 |
| Region groups |  |
| Canada | -1.45 (-2.45, -0.44) |
| West (U.S.) | -0.34 (-0.51, -0.18) |
| South (U.S.) | -0.19 (-0.33, -0.05) |
| Northeast (U.S.) | -0.40 (-0.65, -0.15) |
| Midwest (U.S.) | -0.48 (-0.72, -0.24) |
| Test for subgroup differences (*p*) | 0.0353 |

**Figure 2.**
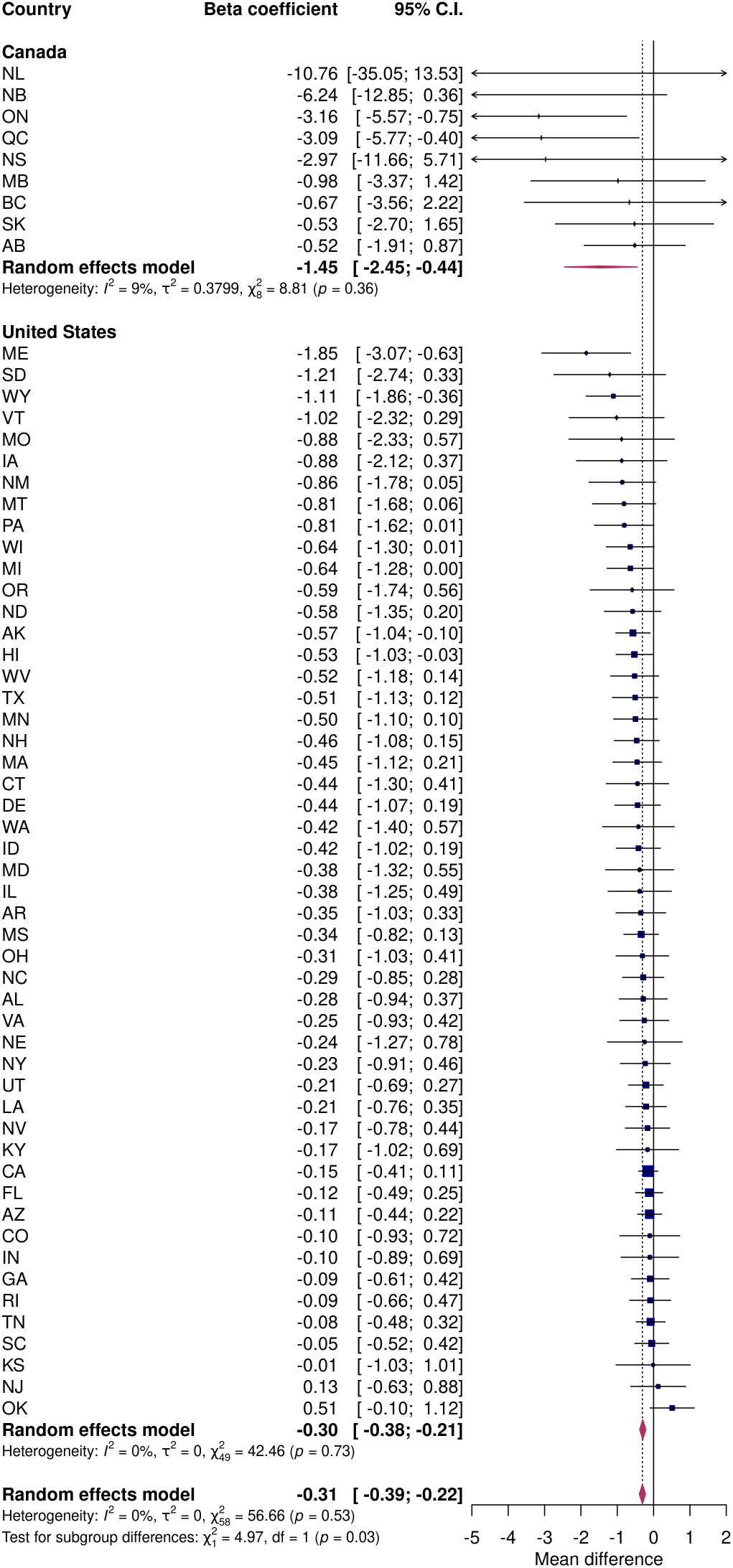
Subgroup meta-analysis by country of the responsiveness of mobility to reported COVID-19 case rates when cases are rising (*β*_*rising*_).

**Figure 3.**
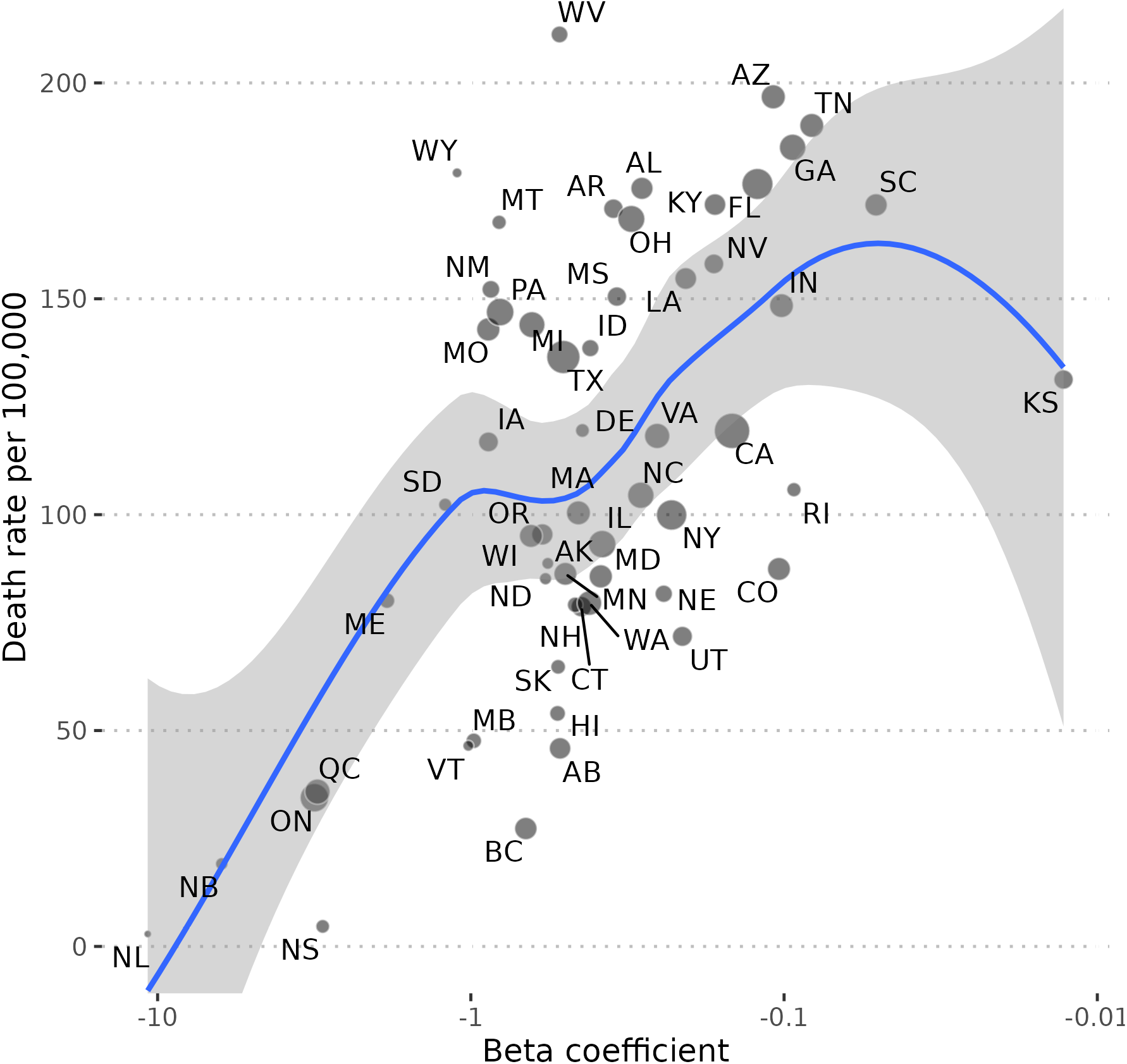
Association between the responsiveness of mobility to reported COVID-19 case rates when cases are rising (*β*_*rising*_) and the rate of COVID-19 deaths from December 2020 to November 2021. Beta coefficients are plotted on a log scale and a loess curve with 95% confidence interval is shown. Point size is proportional to the square root of a jurisdiction’s population. The jurisdictions of New Jersey and Oklahoma are omitted because their beta coefficients had positive signs. Deaths were shifted back by four weeks to account for delays between infections and health outcomes as well as reporting delays.

Responsiveness when cases were falling (*β*_*falling*_) was smaller in magnitude and differed less between countries (Canada: -0·88; 95% CI: -2·00, 0·24 and United States: -0·22; 95% CI: -0·31, -0·13) (table S1). Responsiveness when cases were falling was weakly associated with COVID-19 death rates during the study period (Spearman’s *ρ* = 0·13).

Average mobility was weakly associated with COVID-19 death rates during the study period (Spearman’s *ρ* = 0·20) (figure S1).

The overall estimate of *β*_*rising*_ was attenuated when the lag between mobility and reported case rate was increased from one week (-0·31; 95% CI: -0·39, -0·22) to two (-0·17; 95% CI: -0·26, -0·08) or three (-0·17; 95% CI: -0·25, -0·08) weeks. The estimate of *β*_*falling*_ at one week (-0·23; 95% CI: -0·32, -0·14) differed at two (-0·25; 95% CI: -0·36, -0·14) and three (-0·07; 95% CI: -0·16, 0·03) weeks.

In the alternative interaction-only model, we observed the same patterns to the data as in the primary model (table S2, figure S2). Responsiveness when cases were rising was associated with COVID-19 death rates during the study period (Spearman’s *ρ* = 0·61, figure S3); responsiveness when cases were falling was weakly associated with this outcome (Spearman’s *ρ* = 0·16, respectively).

In the alternative model, the overall estimate of *β*_*rising*_ was attenuated when the lag between mobility and reported case rate was increased from one week (-0·44; 95% CI: -0·51, -0·37) to two (-0·29; 95% CI: -0·36, -0·22) or three (-0·22; 95% CI: -0·29, -0·15) weeks. The estimate of *β*_*falling*_ at one week (-0·14; 95% CI: -0·22, -0·06) was attenuated at two (-0·11; 95% CI: -0·19, -0·03) and three (- 0·01; 95% CI: -0·08, 0·07) weeks.

## Discussion

In this study, we demonstrated a negative relationship between reported COVID-19 incidence and changes in population-level mobility in the subsequent week across nearly every state and province of the United States and Canada. The magnitude of this responsiveness of mobility to COVID-19 cases was nearly five time greater in Canada compared to the United States. Additionally, greater responsiveness was correlated with lower COVID-19 death rates during the study period (December 2020 to November 2021).

Much has been written about the use of mobility data for surveillance and policymaking during the COVID-19 pandemic, showing its utility for evaluating non-pharmaceutical interventions and for short-term forecasting of cases, hospitalizations, and deaths.^1,3–6,9,10,12^ However, our analysis examined the opposite side of the coin: how different jurisdictions reacted (in terms of mobility) to rising case rates. In highly responsive jurisdictions, rising case rates were met by substantial reductions in mobility; in less responsive jurisdictions, only small reductions in mobility were observed for the same levels of incidence.

Reductions in mobility in response to rising case rates are achieved through a mixture of top-down policies and bottom-up individual actions. Risk mitigation behaviour in response to perceived risk has been formalized in the context of sexually transmitted infections^18^ and was explored in relation to the COVID-19 pandemic by Besley and Dray^19^ in their study of the intensity of social distancing (measured using a mobile phone data-derived index) in U.S. counties from January 2020 to January 2021. The authors found that the intensity of social distancing increased with the number of COVID-19 cases (and deaths) reported in the last 7 days. This effect was stronger in the presence of formal restrictions (lockdowns/shop closures) but remained in their absence, pointing to individual responses. They also found that the responsiveness of social distancing to case levels was greater in counties that were Democrat-leaning, high income, more educated, and had lower health insurance coverage.

While our analysis revealed that mobility was responsive to case rates to some degree in nearly every jurisdiction, the subgroup meta-analysis highlighted the pronounced differences in the public health responses between Canada and the United States, with Canadian provinces being nearly five times more responsive than U.S. states on average. This observation comports with broader comparisons between the two countries: Canada recorded much lower rates of COVID-19 cases and mortality than the United States.^20^ Importantly, Canada also recorded much lower maximum values for COVID-19 incidence: in highly responsive jurisdictions, the very high incidence rates seen in many U.S. states were not observed (figure 1).

The strongest measures of responsiveness were seen in Canada’s Atlantic provinces, which adopted an elimination strategy during the study period.^21^ Atlantic Canada saw much lower maximum COVID-19 case rates than the other Canadian provinces (figure S4) and recorded extremely low case rates throughout much of the study period. Atlantic Canada achieved high responsiveness by implementing strong public health measures following upticks in COVID-19 incidence that would be considered unremarkable in any other jurisdiction, in addition to adopting measures to prevent importation.^21^

Limitations exist in the datasets we used in our analysis. We treated reported COVID-19 case and death data as comparable across the jurisdictions included in our study, despite differences in ascertainment and reporting between jurisdictions. For example, the United States had a higher testing rate than Canada.^22^ However, this reflects a truly greater underlying burden of infection: estimates of infection-derived seroprevalence in Canada in November 2021 were generally under 10%^23^; the comparable estimate for United States was around 30%.^24^

Mobility from the Google Community Mobility Reports reflect a subset of the population with mobile devices signed into Google and opted into the “location history” feature. Nonetheless, we believe this measure to be minimally biased, as it need only be representative of the *trends* in mobility in the general population. A weakness in this dataset is that all data are compared to a baseline in January–February of 2020. While we justify our selection of retail and recreation mobility (over other types of mobility, such as workplace visits and time spent at home) above, there are a few instances where this choice causes issues. For examples, for jurisdictions with heavy winter tourism, such as Florida, this mid-winter baseline period leads to lower-than-expected estimates of retail and recreation mobility for the rest of the year, although trends (and thus changes, which we used in our analysis) should be preserved. These issues notwithstanding, the high elasticity of retail and recreation mobility to pandemic-related measures makes it a valuable proxy for risk of disease transmission.^15^

Reverse causality is another concern in our study. Mobility affects case rates.^5–9^ The reverse arrangement, which we examined in our study, is also obviously true: reported case rates affect risk assessment, leading to policies and behaviour changes to limit transmission by reducing mobility.^19,25^ In agreement with our stated hypothesis, we found that mobility was more responsive to COVID-19 incidence when cases were rising (*β*_*rising*_) than when cases were falling (*β*_*falling*_). We expected this because perceived risk should be lower for the same absolute incidence when cases are going up rather than down, resulting in differences in the actions taken to reduce disease transmission. In our sensitivity analysis looking at longer lags between reported case rates and mobility (two or three weeks instead of one) for the calculation of responsiveness, we saw the association diminish as expected, since newer information should have a greater influence on present behaviour.

Additionally, our measure of responsiveness was much more strongly correlated to COVID-19 deaths reported during the study period than was average mobility (Spearman’s *ρ* = 0·51 and 0·20, respectively). Collectively, these findings suggest our responsiveness metric captures something meaningful beyond just the expected correlation between mobility and SARS-CoV-2 transmission.

Cell phone mobility data has proven itself invaluable as a proxy for contact rates and tracking the risk of spread of SARS-CoV-2. In this analysis, we have shown how mobility data may be used to characterize how efficiently various jurisdictions responded to this pandemic threat using a novel metric of the responsiveness of human mobility to rising case rates. We hope our work serves to emphasize the value of this fledging data source for infectious disease surveillance and to highlight that jurisdictions that responded to the pandemic in a vigorous and dynamic way ultimately saw better health outcomes, at least in the near term. In dealing with present and future infectious disease threats, in the words of Dr. Michael Ryan, “the greatest error is not to move.”^26^

## Data Availability

Data and code to reproduce the analysis are available at: https://github.com/jeanpaulrsoucy/covid-19-mobility-responsiveness.

## Author contributions

J-PRS and KAB conceived of the study. J-PRS performed the analysis and wrote the initial draft. All authors contributed to and approved the final manuscript.

## Acknowledgements

DNF is supported by funding from the Canadians Institutes for Health Research (2019 COVID-19 rapid researching funding OV4-170360).

## Supplementary Material

### Supplementary methods

#### Reported COVID-19 cases and deaths

The time series of the total number of COVID-19 cases reported for each U.S. state was retrieved from the Centers for Disease Control and Prevention COVID Data Tracker.^1^ The time series of the total number of COVID-19 cases reported for each Canadian province was retrieved from the Public Health Agency of Canada daily epidemiology update.^2^ Time series on reported COVID-19 deaths were retrieved from the same sources.

#### Population estimates

The mid-year population estimates for U.S. states in 2021 were retrieved from the U.S. Census Bureau.^3^ The mid-year population estimates for Canadian provinces in 2021 were retrieved from Statistics Canada.^4^

**Table S1.** Pooled beta coefficients for the association between weekly COVID-19 case rates and changes in mobility in the subsequent week when cases are falling (*β*_*falling*_) from subgroup meta-analysis by country and region groups.

| **Subgroup** | **Beta coefficient (95% CI)** |
| --- | --- |
| Overall | -0.23 (-0.32, -0.14) |
| Countries |  |
| Canada | -0.88 (-2.00, 0.24) |
| United States | -0.22 (-0.31, -0.13) |
| Test for subgroup differences (*p*) | 0.2475 |
| Region groups |  |
| Canada | -0.88 (-2.00, 0.24) |
| West (U.S.) | -0.32 (-0.52, -0.13) |
| South (U.S.) | -0.13 (-0.28, 0.02) |
| Northeast (U.S.) | -0.27 (-0.55, 0.01) |
| Midwest (U.S.) | -0.27 (-0.51, -0.03) |
| Test for subgroup differences (*p*) | 0.3809 |

**Table S2.** Pooled beta coefficients for the association between weekly COVID-19 case rates and changes in mobility in the subsequent week from subgroup meta-analysis by country and region groups for the interaction-only model.

| Subgroup | Beta coefficient (95% CI) |  |
| --- | --- | --- |
| | Cases rising ($\beta_{rising}$) | Cases falling ($\beta_{falling}$) |
| Overall | -0.44 (-0.51, -0.37) | -0.14 (-0.22, -0.06) |
| Countries |  |  |
| Canada | -1.81 (-2.72, -0.91) | -0.49 (-1.50, 0.53) |
| United States | -0.42 (-0.49, -0.35) | -0.13 (-0.21, -0.05) |
| Test for subgroup differences (*p*) | 0.0028 | 0.4947 |
| Region groups |  |  |
| Canada | -1.81 (-2.72, -0.91) | -0.49 (-1.50, 0.53) |
| West (U.S.) | -0.45 (-0.60, -0.30) | -0.30 (-0.50, -0.09) |
| South (U.S.) | -0.37 (-0.48, -0.26) | 0.00 (-0.13, 0.14) |
| Northeast (U.S.) | -0.54 (-0.74, -0.34) | -0.15 (-0.38, 0.09) |
| Midwest (U.S.) | -0.54 (-0.73, -0.35) | -0.22 (-0.43, -0.02) |
| Test for subgroup differences (*p*) | 0.0151 | 0.0977 |

**Figure S1.**
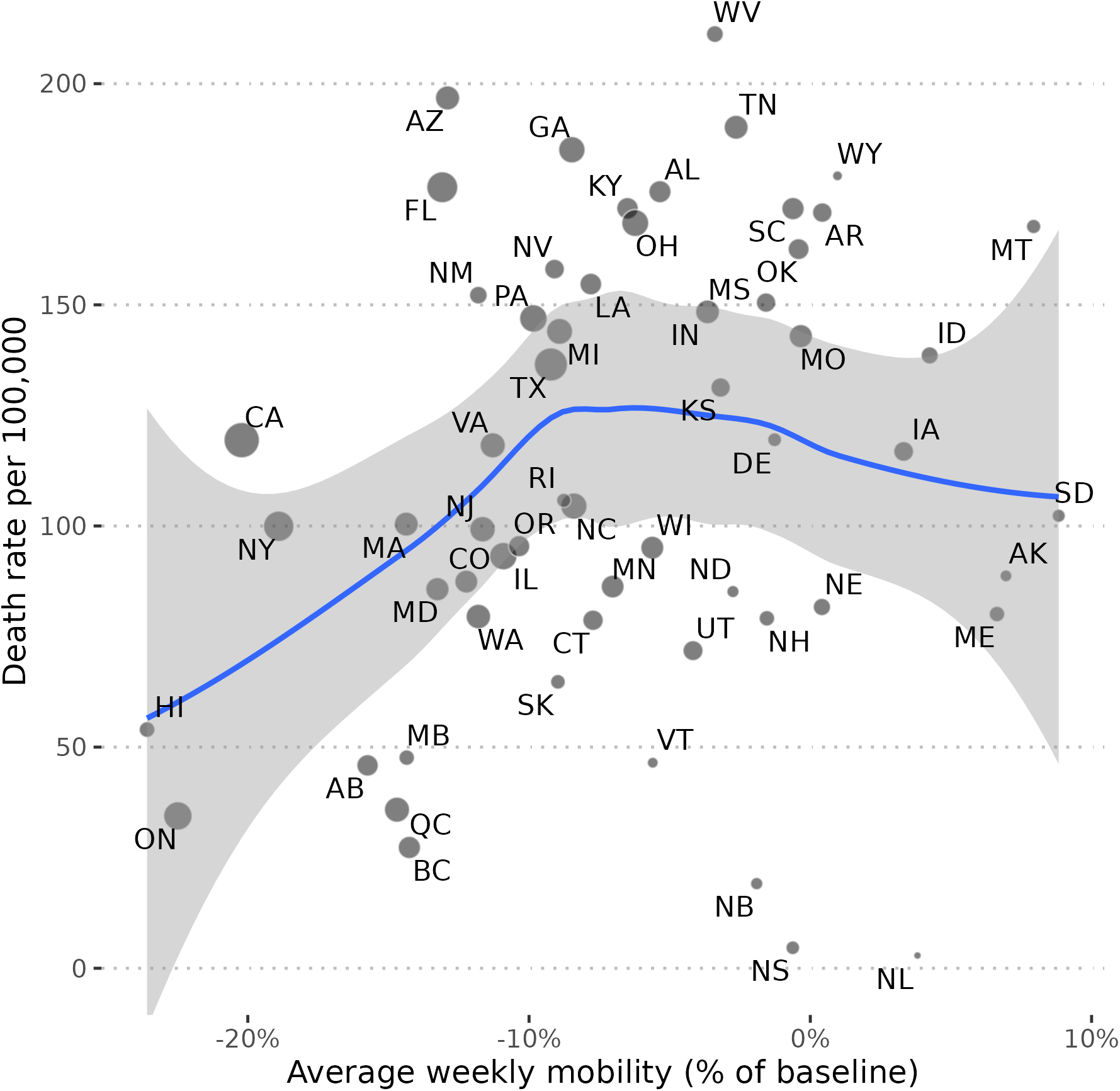
Association between average weekly mobility and the rate of COVID-19 deaths from December 2020 to November 2021. A loess curve with 95% confidence interval is shown. Point size is proportional to the square root of a jurisdiction’s population. Deaths were shifted back by four weeks to account for delays between infections and health outcomes as well as reporting delays.

**Figure S2.**
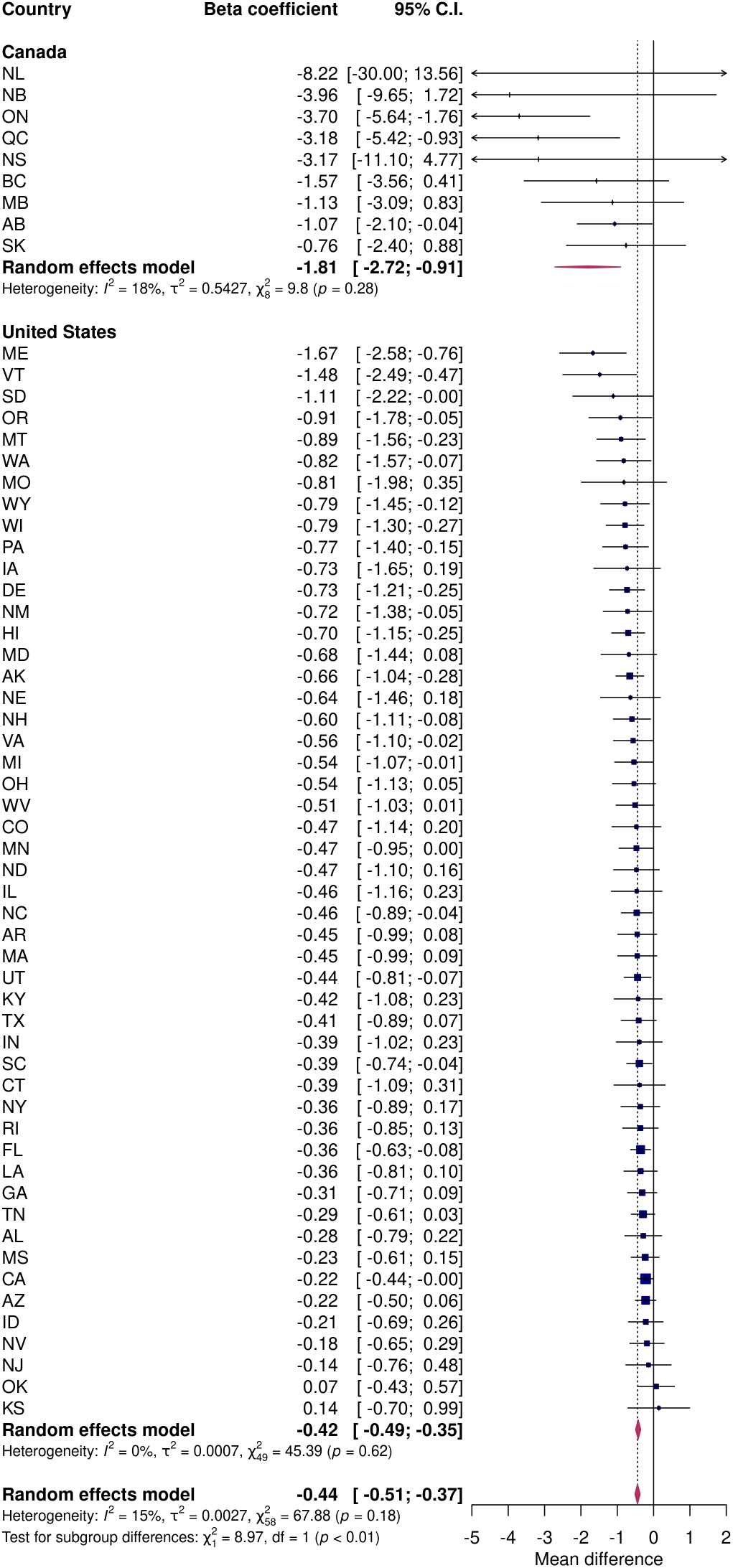
Subgroup meta-analysis by country of the responsiveness of mobility to reported COVID-19 case rates when cases are rising (*β*_*rising*_) for the interaction-only model.

**Figure S3.**
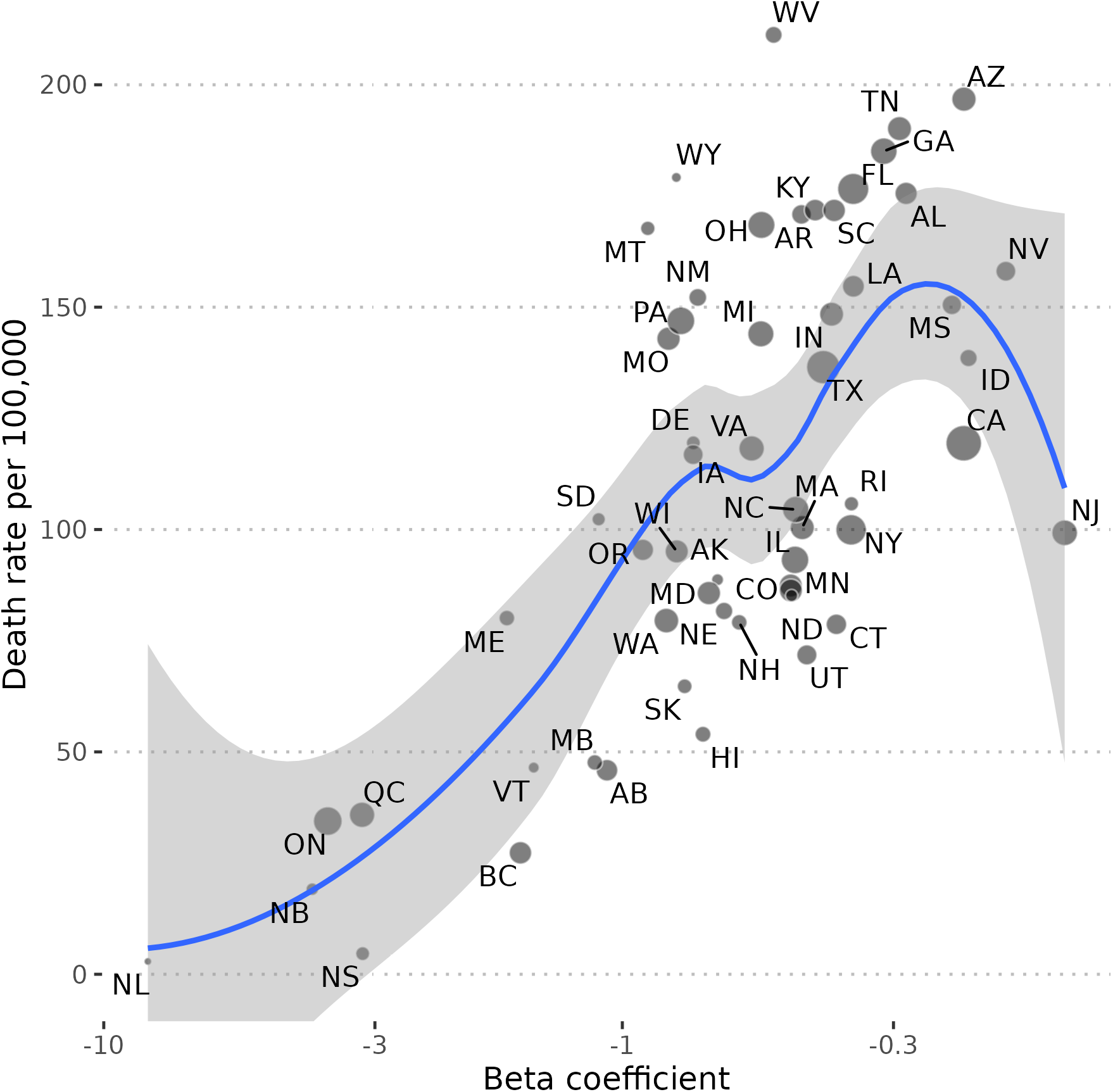
Association between the responsiveness of mobility to reported COVID-19 case rates when cases are rising (*β*_*rising*_) and the rate of COVID-19 deaths from December 2020 to November 2021 for the interaction-only model. Beta coefficients are plotted on a log scale and a loess curve with 95% confidence interval is shown. Point size is proportional to the square root of a jurisdiction’s population. The jurisdictions of Kansas and Oklahoma are omitted because their beta coefficients had positive signs. Deaths were shifted back by four weeks to account for delays between infections and health outcomes as well as reporting delays.

**Figure S4.**
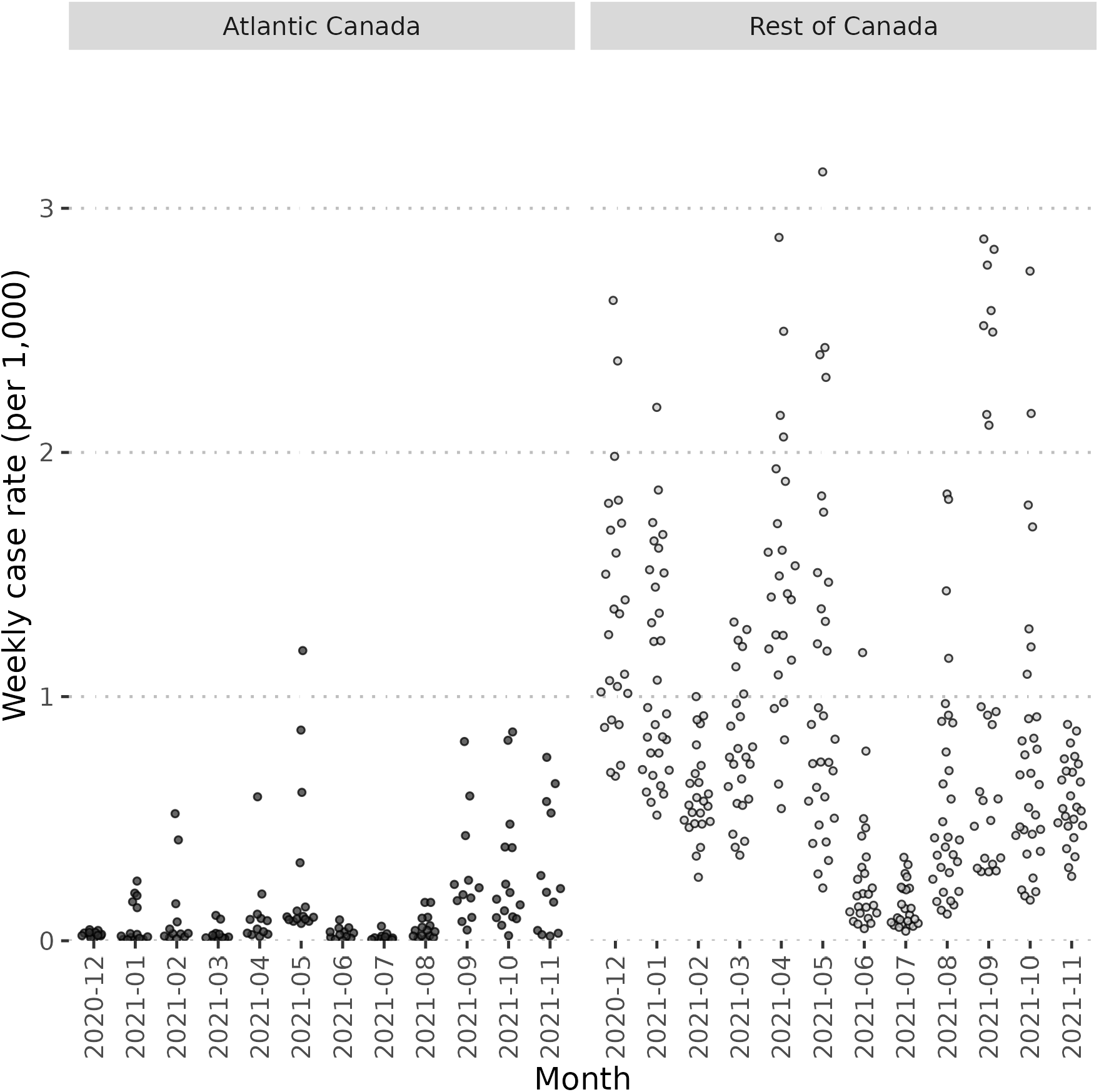
Beeswarm plot of weekly reported COVID-19 case rates per 1,000 population between December 2020 and November 2021 for three provinces in Atlantic Canada (excluding Prince Edward Island) and the remaining six provinces in the rest of Canada.

